# High-throughput *PRPF31* variant characterisation pipeline consistent with ACMG/AMP clinical variant interpretation guidelines

**DOI:** 10.1101/2020.04.06.20055020

**Authors:** Liliya Nazlamova, Man-Kim Cheung, Jelmer Legebeke, Jenny Lord, Reuben J. Pengelly, William Tapper, Gabrielle Wheway

## Abstract

Mutations in *PRPF31* are the second most common cause of the degenerative retinal condition autosomal dominant retinitis pigmentosa. Difficulty in characterising missense variants in this gene presents a significant challenge in providing accurate diagnosis for patients to enable targeted testing of other family members, aid family planning, allow pre-implantation diagnosis and inform eligibility for gene therapy trials. With PRPF31 gene therapy in development, there is an urgent need for tools for accurate molecular diagnosis. Here we present a high-throughput high content imaging assay providing quantitative measure of effect of missense variants in *PRPF31* which meets the recently published criteria for a baseline standard *in vitro* test for clinical variant interpretation. This assay utilizes a new and well-characterized *PRPF31*^*+/-*^ human retinal cell line generated using CRISPR gene editing, which allows testing of *PRPF31* variants which may be causing disease through either haploinsufficiency or dominant negative effects, or a combination of both. The mutant cells have significantly fewer cilia than wild-type cells, allowing rescue of ciliogenesis with benign or mild variants, but do not totally lack cilia, so dominant negative effects can be observed. The results of the assay provide BS3_supporting evidence to the benign classification of two novel uncharacterized *PRPF31* variants and suggest that one novel uncharacterized *PRPF31* variant may be pathogenic. We hope that this will be a useful tool for clinical characterisation of *PRPF31* variants of unknown significance, and can be extended to variant classification in other ciliopathies.

## 1 Introduction

Ciliopathies are a broad range of inherited developmental and degenerative diseases associated with structural or functional defects in motile or primary non-motile cilia (Oud et al. 2017). Motile ciliopathies, such as primary ciliary dyskinesia, commonly present with severe respiratory problems and *situs* defects. Primary non-motile ciliopathies include both syndromic multi-organ conditions, such as Joubert syndrome and Alström syndrome, as well as single-organ disorders such as polycystic kidney disease and some forms of retinitis pigmentosa and Leber congenital amaurosis which only affect the retina. Common clinical features of these non-motile ciliopathies include retinal degeneration and kidney disease; around one third of all cases of retinal dystrophy can be considered retinal ciliopathies, arising as a result of defects in the photoreceptor cilium. Whilst individually rare, collectively, ciliopathies are estimated to affect ∼1:1000 people in the general population worldwide, affecting ∼67,500 people in the UK (Wheway et al. 2019a). However, this is likely to be an underestimate, as ciliopathies are likely to be under-diagnosed.

Ciliopathies are genetic, mostly autosomal recessive, conditions. There are ∼200 known ciliopathy disease genes and it is expected that there are many more unidentified. Genetic testing can provide an accurate diagnosis, but 24-60% of ciliopathy patients who undergo genetic testing do not receive a genetic diagnosis (Bachmann-Gagescu et al. 2015; Knopp et al. 2015; Sawyer et al. 2016; Watson et al. 2016). This is at least in part due to the fact that following current guidelines from the American College of Medical Genetics (Richards et al. 2015), it is difficult to provide a confident clinical diagnosis of disease caused by missense or non-coding variants, which account for more than one third of cases of disease. It is estimated that around 10% of ciliopathy patients in the UK have plausibly pathogenic missense mutations in known disease genes which cannot be classified as pathogenic following current ACMG guidelines because they lack sufficient supporting evidence (eg segregation, recurrence, splicing etc).

The difficulty in classification stems from the requirement for labour-intensive functional studies, and the lack of clarity in ACMG guidelines as to what constitutes a valid functional assay. Variant Curation Expert Panels (VCEPs) have developed guidelines for valid functional assays for specific conditions, but these vary widely from *in vitro* assays, splicing assays to animal model studies (Kanavy et al. 2019). A recent publication (Brnich et al. 2019) outlines general guidelines for assessing whether *in vitro* assays meet baseline standard for clinical variant interpretation, stating the following criteria:

1. The disease mechanism must be understood
2. Assays must be applicable to this disease and this disease mechanism
3. Normal/negative/wild-type AND abnormal/positive/null controls must be used AND multiple replicates must be used
4. Variant controls must be known benign and known pathogenic
5. Statistical analyses must be applied to calculate the level of evidence for each variant

To facilitate standardized application of levels of evidence, Brnich et al 2019 provide tables for calculating odds of pathogenicity values (OddsPath), with each OddsPath equating to a corresponding level of evidence strength (supporting, moderate, strong, very strong) in keeping with the ACMG/AMP variant interpretation guidelines (Richards et al. 2015). This provides a useful framework for developing variant analysis pipelines, but the work involved in optimizing and carrying out such robust in vitro assays is often beyond the scope of diagnostic labs, which do not possess the time or resources to carry out such assays for all but the most common disease genes. It is important for academic research laboratories to work with clinical diagnostic laboratories to develop robust, reliable variant analysis pipelines which meet these criteria. This is particularly important as increasing volumes of variants of unknown clinical significance are produced by genome sequencing, which is being integrated into the UK National Health Service as a standard clinical service (Wheway and Mitchison 2019).

Recent imaging screens for genes involved in ciliogenesis have demonstrated the power of high content imaging for analysis of cilia gene function (Kim et al. 2010; Roosing et al. 2015; Wheway et al. 2015; Kim et al. 2016). Disturbance of cilia gene function provides a robust binary output (presence/absence of cilia) which is highly amenable to high-throughput analysis via automated imaging and image analysis, and can provide a continuous data readout in the form of percentage of cells with a single cilium. siRNA screens for novel cilia genes and cilia regulators have been highly successful in identifying novel ciliopathy disease genes and ciliary functional modules (Kim et al. 2010; Wheway et al. 2015; Kim et al. 2016). The advent of CRISPR gene editing provides new opportunities for exploiting such imaging approaches for classification of variants of unknown clinical significance.

One group of retinal ciliopathies (cilia-associated diseases specifically affecting the retina) are the forms of retinitis pigmentosa (RP) associated with mutations in pre-mRNA splicing factors *PRPF3, 4, 6, 8, 31, SNRNP200, CWC27* and *RP9*. Collectively these are the second most common cause of autosomal dominant RP. Although it remains unclear why, defects in these pre-mRNA splicing factors lead to a degenerative retinal cilia phenotype which can be observed in cells harbouring pathogenic variants in these genes in the laboratory (Wheway et al. 2015; Buskin et al. 2018; Brydon et al. 2019).

All reported variants in *PRPF3, 4, 6, 8, SNRNP200, CWC27* and *RP9* are missense mutations. Most reported variants in *PRPF31* are null variants (Martin-Merida et al. 2018; Wheway et al. 2020), but there are many missense variants in *PRPF31* in public mutation databases which are labelled ‘uncertain clinical significance’. Mutations in *PRPF31* are the most common cause of autosomal dominant RP after rhodopsin mutations, and characterization of missense variants in this gene presents a significant challenge in providing accurate diagnosis for patients. Developing tools to provide accurate genetic diagnoses in these cases is a significant clinical priority, particularly as *PRPF31* gene therapy is in development (Brydon et al. 2019).

In this study we use CRISPR gene editing and high throughput imaging of ciliated cells to establish a variant analysis pipeline consistent with recommendations for application of the functional evidence PS3/BS3 criterion (PS3 = well-established functional studies show a deleterious effect, BS3 = well-established functional studies show no deleterious effect) using the ACMG/AMP sequence variant interpretation framework, for accurate clinical genetic diagnosis of missense variants in *PRPF31*. We studied all *PRPF31* missense variants currently annotated as ‘unknown clinical significance’ in the ClinVar database of variant interpretations (Landrum et al. 2014; Landrum et al. 2016).

## Methods

### *In silico* splicing analysis

We used Human Splicing Finder v3.1 (Desmet et al. 2009) to identify and predict the effect of each variant on splicing motifs, including the acceptor and donor splice sites, branch point and auxiliary sequences known to enhance or repress splicing. This programme uses 12 different algorithms to make a consensus prediction of the effect of variants on splicing.

### *In silico* predictions of functional effect of missense variants

We used Ensembl variant effect predictor (VEP) (McLaren et al. 2016) to predict the functional impact of each missense variant using SIFT (Ng and Henikoff 2003) and PolyPhen-2 (Adzhubei et al. 2013).

### 3D structural protein analysis

PyMOL v2.3.3 (Schrodinger Ltd) programme was used to model the effect of missense variants in human PRPF31 protein. Missense variants were modelled on PRPF31 in complex with 15.5K (SNU13) and U4 snRNA (PDB file 5o9z) (Liu et al. 2007) and on PRPF31 in the tri-snRNP (PDB file 5o9z) (Bertram et al., 2017). Both of these complexes are part of the pre-catalytic spliceosome.

### *In silico* predictions of pathogenicity

We used CADD to calculate relative estimates of pathogenicity of variants (Rentzsch et al. 2019). Higher raw values indicate higher likelihood of pathogenicity. PHRED corrected values are normalized to all potential SNVs across the genome and again higher values indicate higher likelihood of deleterious effects compared to all other variants.

### CRISPR gene knockouts

*Streptococcus pyogenes* Cas9 (spCas9) was complexed with one of four modified single guide RNAs (sgRNAs) targeting intron 4 or exon 5 of *PRPF31* (Synthego) to form ribonucleoprotein complexes (RNPs). sgRNA sequences were: sgRNA1 TCTGCTCGCCCCCAGGAGCT (PAM GGG), sgRNA2 CATTGTTCTTGCACTTGTCC (PAM AGG), sgRNA3 GACGACCATGATGGTGGCAT (PAM TGG), sgRNA4 AGGGAGGCGCCGGGCCCTAA (PAM TGG). sgRNAs had the following modifications to increase stability: 2’-O-methyl analogs and 3’ phosphorothioate internucleotide linkages at the first three 5’ and 3’ terminal RNA residues. RNPs were prepared in 1:6 (vol:vol) ratio (protein to modified RNA oligonucleotide) in P3 solution (supplemented) and incubated for 10 mins at room temperature prior nucleofecting the cell suspension (100,000 cells/5µl P3 reagent per reaction, Lonza protocol EA104).

### Analysis of on-target changes

A proportion of bulk edited cells were harvested for DNA extraction and PCR amplification of the relevant targeted region of *PRPF31* using OneTaq polymerase (NEB). PCR products were cleaned using ExoSAP-IT (Thermo Fisher) and Sanger sequencing was performed by Source Biosciences. Sequencing traces were analysed using inference of CRISPR edits (ICE) analysis (Synthego). Of the four gRNAs tested, indel frequencies and knockout efficiencies were as follows: guide 1 32%/29%, guide 2 43%/37%, guide 3 40%/26%, guide 4 85%/72%. Cells edited with guide 1 were taken forward for single cell isolation.

### Single cell cloning

Cells were dissociated using Accutase at room temperature, counted and transferred to a conical tube. Cells were collected by centrifugation at 200 g and washed with sterile sort buffer (Ca & Mg free PBS, 25 mM HEPES pH 7.0, 1-2.5 mM EDTA and 0.5% BSA or 1-2% FCS). Cells were collected again and resuspended at a concentration of 5-8×10^6^ cells/ml. Untransfected cells were used for gating cell size on the FACS Aria cell sorter (BD) and edited cells then sorted into 150 µL DMEM/F12 + 20% FCS + 10% antibiotic and antimycotic + 10 µM Y-27632 ROCK inhibitor (STEMCELL Technologies) into each well of a 96 well plate.

### Off-target effect prediction

Cas-OFFinder (Bae et al. 2014) was used to predict potential off-target cut sites of Cas9 guided by sgRNA1. Allowing up to 3 nucleotide mismatches of the sgRNA, 15 potential off-target sites were identified in GRCh38, including 6 in introns, 1 in 3’UTR, 1 in a non-coding exon and 2 at intron/exon boundaries. These regions were visually inspected for insertions or deletions or SNVs in RNA sequence (details below) using Integrative Genomics Viewer (Robinson et al. 2011).

### Cell fractionation

Cells were fractionated into nuclear and cytoplasmic fractions. Cells were collected by scraping into fractionation buffer (20mM HEPES pH7.4, 10mM KCl, 2mM MgCl_2_, 1mM EDTA, 1mM EGTA) on ice, lysed through a 27 gauge needle, on ice. The nuclear pellet was collected by centrifugation at 720 x g, washed and dispersed through a 25 gauge needle. The supernatant containing cytoplasm was centrifuged at 10,000g to remove mitochondria and any cell debris. The dispersed nuclear pellet was collected again by centrifugation at 720 x g, resuspended in TBS with 0.1% SDS and sonicated to shear genomic DNA and homogenize the lysate.

### RNA extraction

RNA was extracted from fractionated samples using TRIzol Reagent (Thermo Fisher). RNA quality and concentration was measured using an RNA Nano chip on the Agilent Bioanalyser 2100. Samples with total RNA concentration ≥20ng/µl, RIN ≥6.8 and OD 260/280 were taken forward for cDNA library preparation and sequencing.

### cDNA library preparation and sequencing

cDNA libraries were prepared using Ribo-Zero Magnetic Kit for rRNA depletion and NEBNext Ultra Directional RNA Library Prep Kit library prep kit by Novogene Inc. Library quality was assessed using a broad range DNA chip on the Agilent Bioanalyser 2100. Library concentration was assessed using Qubit and q-PCR. Libraries were pooled, and paired-end 150bp sequencing to a depth of 20M reads per fraction (40M reads per sample) was performed on an Illumina HiSeq2500 by Novogene Inc.

### Data processing

#### Raw data quality control

Raw FASTQ reads were subjected to adapter trimming and quality filtering (reads containing N > 10%, reads where >50% of read has Qscore<= 5) by Novogene Inc.

Quality of sequence was assessed using FastQC v0.11.5 (https://www.bioinformatics.babraham.ac.uk/projects/fastqc/). No further data filtering or trimming was applied.

#### Data deposition

Raw FASTQ reads after adapter trimming and quality filtering (reads containing N > 10%, reads where >50% of read has Qscore<= 5) were deposited on the Sequence Read Archive, SRA accession PRJNA622794.

#### Alignment for transcript level analysis

Paired FASTQ files were aligned to GRCh38 human genome reference using GENCODE v29 gene annotations (Frankish et al. 2019) and STAR v2.6.0a splice aware aligner (Dobin et al., 2013), using ENCODE recommend options (3.2.2 in the STAR manual (https://github.com/alexdobin/STAR/blob/master/doc/STARmanual.pdf). The two-pass alignment method was used, with soft clipping activated.

#### Alignment quality control and transcript level abundance estimates

BAM files sorted by chromosomal coordinates were assessed for saturation of known splice junctions and transcript abundance estimates in fragments per kilobase of exon per million reads (FPKM) were calculated using RSeqQC v3.0.1 (Wang et al., 2012, Wang et al., 2016).

#### Differential splicing analysis

rMATs v4.0.2 (rMATS turbo) (Shen et al. 2014) was used to statistically measure differences in splicing between replicates of wild-type and mutant sequence. BAM files aligned with STAR v2.6.0a two-pass method with soft clipping suppressed were used as input.

#### Protein extraction

Total protein was extracted from cells using 1% NP40 lysis buffer and scraping. Insoluble material was pelleted by centrifugation at 10,000 x g. Cell fractionation was carried out by scraping cells into fractionation buffer containing 1mM DTT and passed through a syringe 10 times. Nuclei were pelleted at 720 x g for 5 minutes and separated from the cytoplasmic supernatant. Insoluble cytoplasmic material was pelleted using centrifugation at 10,000 x g for 5 minutes. Nuclei were washed, and lysed with 0.1% SDS and sonication. Insoluble nuclear material was pelleted using centrifugation at 10,000 x g for 5 minutes.

#### SDS-PAGE and western blotting

20µg of total protein per sample with 2 x SDS loading buffer was loaded onto pre-cast 4-12% Bis-Tris gels (Life Technologies) alongside Spectra Multicolor Broad range Protein ladder (Thermo Fisher). Samples were separated by electrophoresis. Protein was transferred to PVDF membrane. Membranes were incubated with blocking solution (5% (w/v) non-fat milk/PBS), and incubated with primary antibody overnight at 4°C. After washing, membranes were incubated with secondary antibody for 1 hour at room temperature and exposed using 680nm and/or 780nm laser (LiCor Odyssey, Ferrante, Giorgio et al.), or incubated with SuperSignal West Femto reagent (Pierce) and exposed using Chemiluminescence settings on ChemiDoc MP imaging system (Bio-Rad)

#### Primary antibodies for WB

Mouse anti ß actin clone AC-15. 1:4000. Sigma-Aldrich A1978 Mouse anti-c myc 1:5000 (Sigma) Goat anti-PRPF31 primary antibody 1:1000 (AbNova) Rabbit anti-PRPF31 primary antibody 1:1000 (AbCam) Rabbit anti-PRPF6 primary antibody 1:1000 (Proteintech)

#### Secondary antibodies for WB

Donkey anti mouse 680 1:20,000 (LiCor)

Donkey anti goat 800 1:20,000 (LiCor)(Ferrante et al.)

Donkey anti mouse HRP (Dako)

Donkey anti rabbit HRP (Dako)

#### Variant construct cloning

Full-length, sequence-validated *PRPF31* ORF clone with C-terminal myc tag was obtained from Origene. Single nucleotide variants were introduced using NEB Q5 site-directed mutagenesis kit. The entire wild-type and mutant clone sequence was verified by Sanger sequencing (Source Bioscience).

#### Cell culture

hTERT-RPE1 cells (ATCC CRL-4000) were cultured in DMEM/F12 (50:50 mix) + 10% FCS at 37°C, 5% CO_2_, and split at a ratio of 1:8 once per week.

#### Cell transfection

The construct was transfected into hTERT-RPE1 cells using the Lonza 4D Nucleofector. Construct was mixed with P3 solution (supplemented) and incubated for 10 mins at room temperature prior to nucleofecting the cell suspension (100,000 cells/5µl P3 reagent per reaction, Lonza protocol EA104).

#### Imaging plate setup

20µl nucleofected cells were plated at a density of 1 × 10^5^ cells ml^-1^ into 80µl complete media per well in 96 well optical bottom Perkin Elmer ViewPlates. The outer wells were filled with media without cells to reduce edge effects. Cells were cultured for 48 hours before media was changed to serum-free media. Cells were fixed 24 hours later.

#### Immunocytochemistry of imaging plates

Wells were emptied by inversion of plates, and washed with warm Dulbecco’s PBS (Sigma). DPBS was removed by plate inversion and cells were fixed with ice cold methanol for 5 minutes at −80°C. Methanol was removed by plate inversion and cells were washed twice with PBS and non-specific antibody binding sites blocked with 1% non-fat milk powder/PBS (w/v) for 15 minutes at room temperature. Cells were incubated with primary antibodies in blocking solution for 1 hour at room temperature and secondary antibodies + DAPI for 1 hours at room temperature in the dark. Mowiol was added to wells, and plates stored until imaging.

#### Primary antibodies for immunocytochemistry

Rabbit anti-Arl13b primary antibody 1:200 (Proteintech)

Mouse anti-c myc 1:1000 (Sigma)

#### Secondary antibodies for immunocytochemistry

Donkey anti mouse IgG AlexaFluor 488 1:500 (ThermoFisher)

Donkey anti goat IgG AlexaFluor 568 1:500 (ThermoFisher)

#### High-throughput confocal imaging

Imaging was carried out on a Perkin Elmer Opera LX with 20x and 60x water immersion lenses at Wolfson Bioimaging Centre, University of Bristol.

#### Image analysis

Image analysis was performed using custom scripts optimized on CellProfiler (Carpenter et al. 2006). Analysis included nuclear recognition and counting, cell recognition, exclusion of border objects and counting of whole cells, cilia recognition and counting, and quantification of the percentage of whole cells with a single cilium. Median and median absolute deviation of untransfected cells were used to calculate robust z scores (Zhang 2007; Chung et al. 2008; Birmingham et al. 2009) of cell number and percentage of whole cells with a single cilium in transfected cells.

## Results

### *In silico* splicing analysis of genetic single nucleotide variants in *PRPF31*

To exclude any variants annotated as missenses which are actually altering splicing rather than simply encoding missense amino acids (as reported in Rio Frio *et al.*, 2008) we used Human Splicing Finder v3.1 (Desmet et al. 2009)to assess all published missense variants in *PRPF31* and those in ClinVar. Of the twelve published variants in *PRPF31* annotated as missense, five were predicted to potentially alter splicing, and one (c.1373A>T, p. Gln458Leu (Xiao et al. 2017) was predicted to be highly likely to affect splicing, suggesting it may need to be reclassified as a splicing variant (**Table 1**). Of the 39 variants in *PRPF31* labelled ‘uncertain significance’ in ClinVar, 27 are single nucleotide changes in the exons or exon/intron boundaries of the gene. One of these (c.420G>C p.Lys140Asn) is highly likely to affect splicing, as it breaks a wild-type consensus splice donor site, and can be considered probably pathogenic (**Table 1**). Thirteen others potentially affect splicing and can be considered possibly pathogenic (**Table 1**). Functional studies would be needed to clarify effects of these, as ACMG guidelines only allows *in silico* splice tools as supporting evidence of pathogenicity (Richards et al. 2015), but this is beyond the scope of this particular investigation. To avoid investigating any variants which potentially affect splicing, we excluded all variants predicted by Of the thirteen which are not predicted to affect splicing, ten are either within introns, or are exonic but do not result in amino acid changes, and can therefore be considered probably benign (**Table 1**).

### *In silico* predictions of functional effect of missense variants

We used Ensembl variant effect predictor (VEP) to annotate missense variants using SIFT and PolyPhen-2. All previously reported pathogenic variants were predicted deleterious/probably damaging. Of the 15 variants in *PRPF31* labelled ‘uncertain significance’ in ClinVar which are in the exons, 5 were predicted tolerated/benign, 6 were predicted deleterious/probably damaging, 4 were predicted deleterious/possibly damaging and 1 had conflicting predictions (deleterious/benign).

Of these 15 variants only 3 were predicted not to affect splicing, and so we concentrated on characterizing these variants. These were Thr50Ile (deleterious/benign), Met212Val (deleterious/probably damaging), Val433Ile (tolerated/benign).

### 3D structural analysis of missense variants in PRPF31

We have previously mapped all published missense variants onto the PRPF31 protein structure in complex with U4 snRNA and 15.5K (SNU13) protein (Bertram et al. 2017) and shown that variants are located throughout the protein, but concentrated in several key domains; in α-helix 12 of the protein, in the Nop domain which interacts with RNA and the 15.5K (SNU13) protein, in α-helix 1 or α-helix 6 of the coiled-coil domain and α-helix 3 of the protein in the coiled-coil tip(Wheway et al. 2019b). Single pathogenic variants are also found within the flexible loop between the Nop and coiled-coil domains and in the C-terminal domain. In most cases missense substitutions affect hydrogen (H) bonding within PRPF31 or with neighbouring PRPF6 or PRPF8 or introduce a new kink in the amino acid chain, resulting in the loss of a polar contact (Wheway et al. 2019b).

We mapped the three uncharacterised ClinVar missense variants; Thr50Ile, Met212Val, Val433Ile onto the PRPF31 protein structure in complex with U4 snRNA and 15.5K (SNU13) protein (**Figure 1a)**. Thr50Ile is in the unstructured N-terminal domain, Met212Val is in α-helix 6 of the coiled-coil domain and Val433Ile is close to the RNA binding site. Met212Val affects H bonding in PRPF31 (**Figure 1b**) and Thr50Ile affects H bonding between PRPF31 and PRPF6 (**Figure 1c**), and we predict that these variants affect protein folding and solubility, and are pathogenic. Val433Ile does not affect H bonding within PRPF31, between PRPF31 and U4, nor does it affect polar contacts (**Figure 1d**), so we predict this variant to be benign.

**Figure 1.**
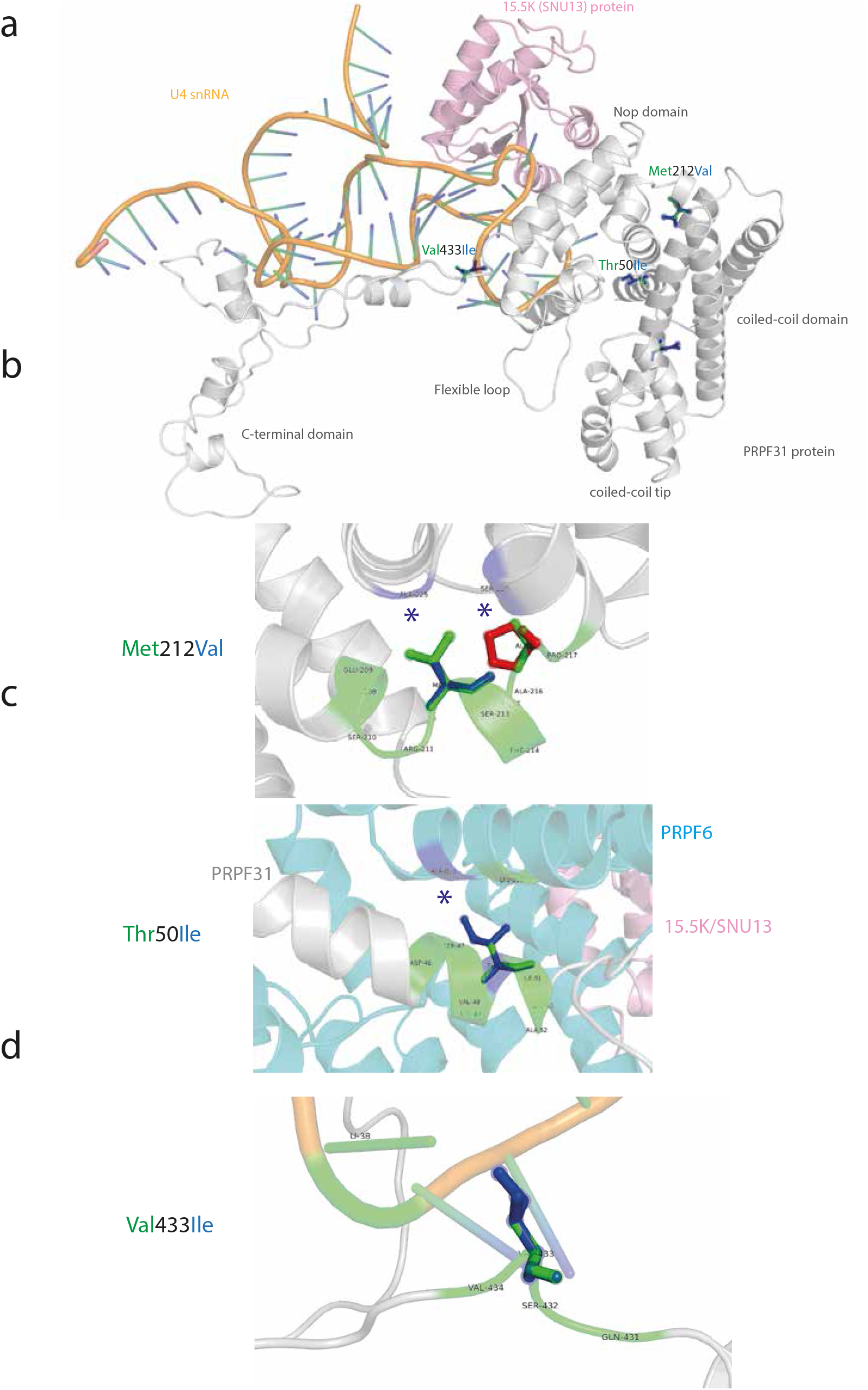
3D cartoon representation of PRPF31, including selected ClinVar missense mutations, and their interactions within 4Å. (a) Cartoon representation of alpha helical structure of PRPF31 (grey) and 15.5K/SNU13 (pink) with U4 snRNA (orange backbone), with ClinVar missense mutations of uncertain significance mapped onto the physical structure, with wild-type amino acid structure in green, and mutant amino acid structure overlaid in blue. (b) Cartoon representation of alpha helical structure of regions of PRPF31 (grey), with published missense mutations of Met212Val, (c) Thr50Ile and (d) Val433Ile mapped onto the physical structure, with wild-type amino acid structure in green, and mutant amino acid structure overlaid in blue, and interactions within 4Å, predicted to affect H bonding within PRPF31 (b-d) or between PRPF31 and PRPF6 (blue) (c). Blue asterisks are used to label where missense mutations introduce new H bonding.

We took these variants, with one negative control and four positive control variants, three of which have been proven to affect PRPF31 protein function in *in vitro* assays, forward for *in vitro* analysis.

### Production and characterisation of *PRPF31* knockout (KO) retinal pigment epithelium (RPE1) cell line

It remains unclear whether missense variants in *PRPF31* cause disease by dominant negative effects or haploinsufficiency. It has been suggested that *PRPF31*-associated disease is caused by a combined dominant negative and haploinsufficiency mechanism (Rose and Bhattacharya 2016; Wheway et al. 2020). In order to produce a disease-relevant human cell model which would allow analysis of PRPF31 variants acting via a mechanism of dominant negative effects or haploinsufficiency, we produced stable monoclonal *PRPF31* heterozygous mutant retinal pigment epithelium (RPE1) cell lines. We achieved this using purified wild-type Cas9 and four single guide RNAs targeting intron 5 and exon 6 (coding exon 5) of *PRPF31* which were modified to increase stability (**Figure 2a**). We achieved up to 85% indel frequency, with up to 72% overall knockout efficiency. From the pool of edited cells from sgRNA1 we used single cell sorting to isolate clones of *PRPF31*^*+/-*^ cells with heterozygous knockouts and wild-type unedited clones. We took three of each on for further analysis. We confirmed insertion of T at the intron 5/exon 6 boundary of *PRPF31* which causes a frameshift and premature termination codon (**Figure 2b**). We performed whole transcriptome sequencing on RNA from the nucleus (a mixture of completely and incompletely spliced transcripts) and cytoplasm (only completely spliced transcripts) from all 6 clones (SRA accession PRJNA622794). We analysed predicted off-target changes in each clone through manual analysis of target regions in our RNAseq data in IGV, via analysis of differential gene expression using the edgeR package (Robinson et al. 2010; McCarthy et al. 2012) and analysis of differential splicing using rMATS turbo (Shen et al. 2014) (**Supplementary Table 2**). We found no evidence of sequence changes or expression changes in any of the genes predicted to be off-target sites (with 3 mismatches) but found statistically significant differential usage of 3 exons in *MEGF6* between wild-type and mutant clones. Exons 25 (ENSE00001477187) and 24 (ENSE00001477188) of ENST00000356575.9, and exon 27 (ENSE00001308186) of ENST00000294599.8 are each significantly skipped in mutants, FDR p value = 0.0279, 0.0343 and 0.0086 respectively) (**Supplementary Table 2**). *MEGF6* is a poorly characterised protein which has not been linked to cilia, and we do not expect this change to affect our cell phenotype, but it is important to note this splicing variation in a gene which could potentially be an off-target effect of our CRISPR guide RNAs. Analysis of splicing patterns of *PRPF31* showed no significant change in splicing of intron 5 or exon 6 in the mutant clones compare to wild-type (no differential 3’ splice site usage, skipping of exon 6 or retention of intron 5-6). However, we did unexpectedly observe an increase of retention of intron 12-13 in the nuclear fraction of the mutant cells (FDR p value = 0.0141 when considering only reads mapping splice junctions, or FDR p value = 0.0091 when also considering reads mapping to the intron), although this was not observed in the cytoplasmic fraction of the cells (**Supplementary Figure 1**). We hypothesise that mutant *PRPF31* may experience changes in the dynamic of splicing, with less efficient removal of introns before export from the nucleus.

**Figure 2.**
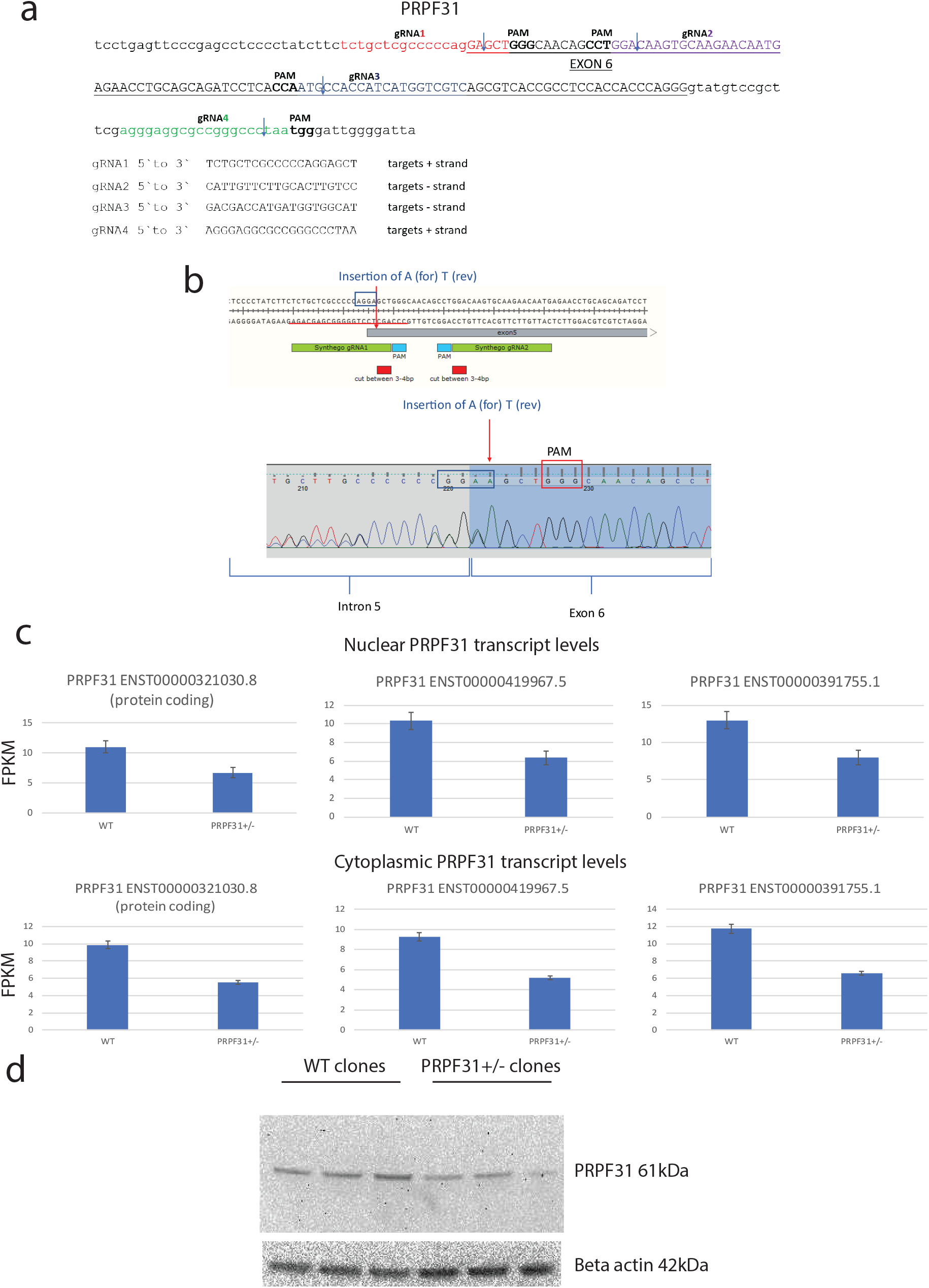
Heterozygous knockout of *PRPF31* in hTERT-RPE1 cells by insertion of single nucleotide in exon 6 by CRISPR/Cas9 editing. (a) Mapping of single guide RNAs to *PRPF31* exon 6 (coding exon 5) used in CRISPR editing approach. (b) Schematic diagram and electropherogram sequence trace showing heterozygous insertion of A near intron 5/exon 6 boundary of *PRPF31* in hTERT-RPE1 cells. (c) Graphs showing roughly 50% reduction in three major *PRPF31* transcripts in edited cells compared to wild-type cells (FPKM = fragments per kilobase of transcript per million mapped reads).* = p <0.05 two-sample t-test (d) western blot showing reduced expression of PRPF31 protein (top) relative to beta-actin control expression (bottom) in edited clones compared to wild-type clones.

Transcript level expression analysis of RNA sequence data showed expression of three *PRPF31* transcripts in both mutant and wild-type cell lines; ENST00000419967.5, ENST00000391755.1 and protein-coding ENST00000321030.8, with an approximately 50% reduction in all *PRPF31* transcripts in the mutant clones (**Figure 2c**). Analysis of reads around the CRISPR insertion site (i.e. at the intron 5/exon 6 boundary) in the mutant clones showed that very few reads contained the insertion. In nuclear RNA from the mutant clones, the ratio of wild-type reads to reads with the insertion was 46:2 (4.2% insertion), 92:11 (10.7% insertion) and 48:0 (0% insertion). Roughly the same proportions of reads with insert were seen in the cytoplasmic RNA from mutant clones (70:2, 61:4, 53:2 ie 2.8%, 6.2%, 3.6%). This suggests that *PRPF31* is preferentially expressed from the wild-type allele in the mutant cells, and both wild-type and mutant transcripts are exported to the cytoplasm. This suggests that in this cell model the disease phenotypes (see later) are caused by haploinsufficiency. Indeed, western blotting of protein extracts from wild-type and mutant clones confirmed reduction in PRPF31 protein levels in mutant cells compared to wild-type control cells with no detectable expression of any mutant protein (**Figure 2d**).

As has been previously reported, mutation of PRPF31 is associated with reduction in the number and length of primary cilia on multiple cell types (Wheway et al., 2015, Buskin et al., 2019, Wheway et al., 2019). To investigate whether this phenotype was observed in our mutant clones in an unbiased way, we used high-throughput imaging and automated image analysis to quantify number of cilia in mutant cells compared to wild-type cells (**Figure 3a**). We also assayed a range of other phenotypes which have been reported in PRPF31 mutants, including cell number, number of micronuclei per cell, nuclear area, nuclear shape (compactness, eccentricity), nuclei staining intensity. Whilst these assays showed a general trend in reduced cell number, increased number of micronuclei per cell and reduced nuclear area in mutant clones compared to wild-type clones, the most robust and consistent phenotype we observed was the loss of cilia phenotype in PRPF31^+/-^ cells (**Figure 3b**).

**Figure 3.**
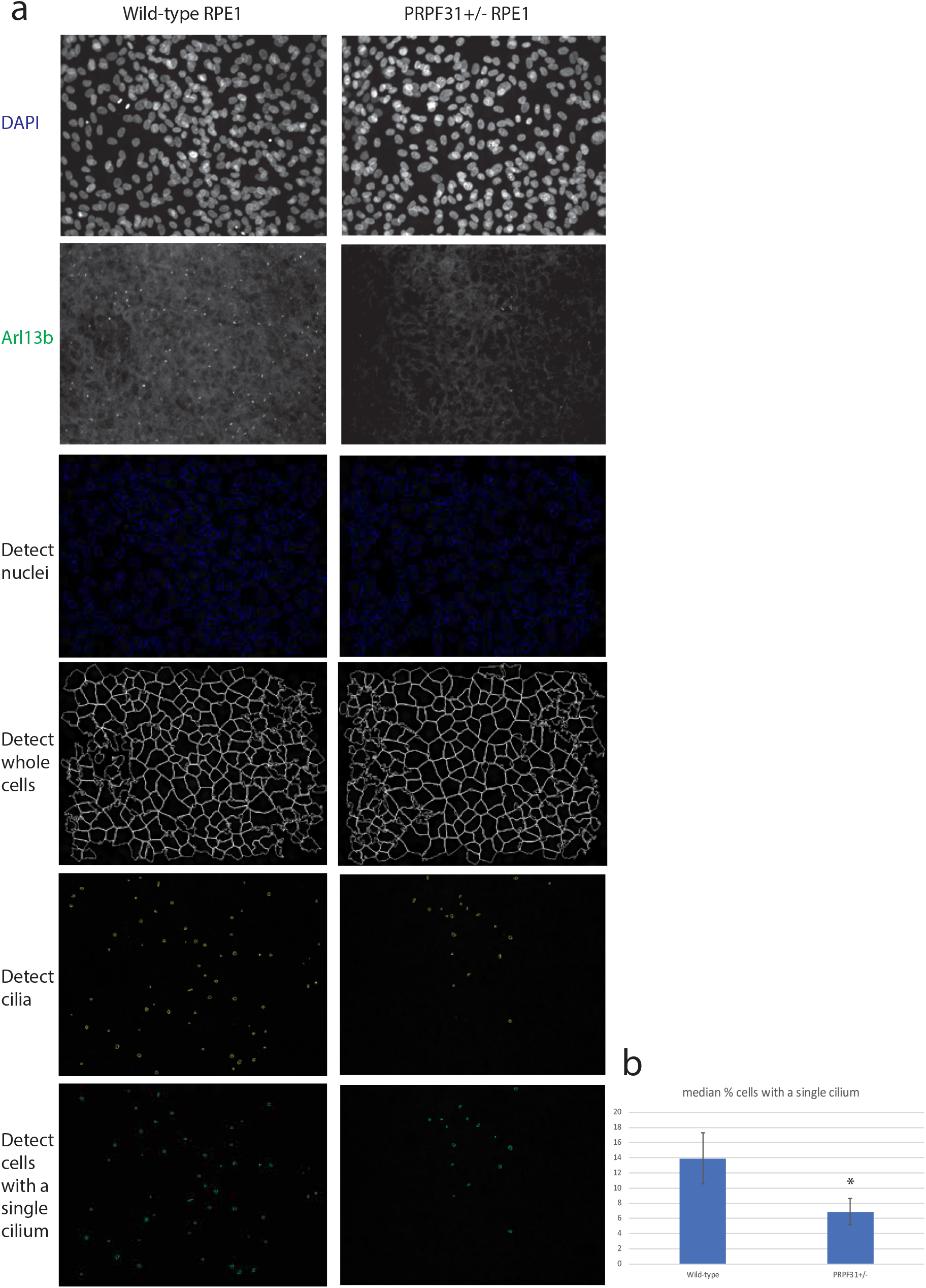
High content image analysis of nuclei and cilia in wild-type and *PRPF31*^*+/-*^ mutant clones. (a) Top two rows of panels show DAPI stained nuclei and Arl13b antibody-stained cilia from wild-type and mutant cells in raw output images from Opera confocal high-throughput imager. Lower four panels show automated image analysis using CellProfiler. (b) Graph showing reduction in the median percentage of cells with a single cilium in edited cells compared to wild-type cells. Error bars = median absolute deviation. * robust z score = 2.73.

### Characterisation of PRPF31 missense variants using high-throughput imaging

We transfected PRPF3^1+/-^ cells with plasmid constructs expressing full-length human PRPF31 with a myc-DDK tag, under the control of a CMV promoter, with various missense mutations introduced by site-directed mutagenesis;

Wild type control PRPF31 WT

Pathogenic control PRPF31:

PRPF31 c.341T>A p.Ile114Asn

PRPF31 c.413C>A p.Thr138Lys

PRPF31 c.581C>A p.Ala194Glu

PRPF31 c.646G>C p.Ala216Pro

And our three variants for testing:

PRPF31 c.149C>T p.Thr50Ile

PRPF31 c.634A>G p.Met212Val

PRPF31 c.1297G>A p.Val433Ile

To investigate their ability to restore cilia growth in the mutant cell line.

We first optimized the assay with positive and negative control variant constructs. To satisfy the requirements of Brnich et al., we included multiple replicates of each construct per plate (3) and repeated each experimental plate in 2 independent replicates. In each well, 6 fields of view were imaged. In each well, the median % cells with a single cilium was measured, and robust z score calculated, comparing this median to the median and median absolute deviation of mock transfected cells(Huang da et al. 2009).

Whilst wild-type PRPF31 rescued the loss of cilia phenotype, the four positive controls (PRPF31 c.341T>A p.Ile114Asn, c.413C>A p.Thr138Lys, c.581C>A p.Ala194Glu and c.646G>C p.Ala216Pro) did not. However, c.413C>A p.Thr138Lys showed more ability to rescue the phenotype than the other controls, suggesting that this is a less severe missense mutation, and according to the recommendations for application of the functional evidence PS3/BS3 criterion using the ACMG/AMP sequence variant interpretation framework, we consider this an intermediate result. As a result we use the Odds of Pathogenicity (OddsPath) estimated by performance of classified variant controls, permitting 1 variant control (either benign or pathogenic) to have an indeterminate readout. The most severe missense mutation was PRPF31 c.581C>A p.Ala194Glu which actually reduced the percentage of cells with a single cilium in the mutant cell line, suggesting that this particular missense mutation has a dominant negative effect on cells (**Figure 4a**).

**Figure 4.**
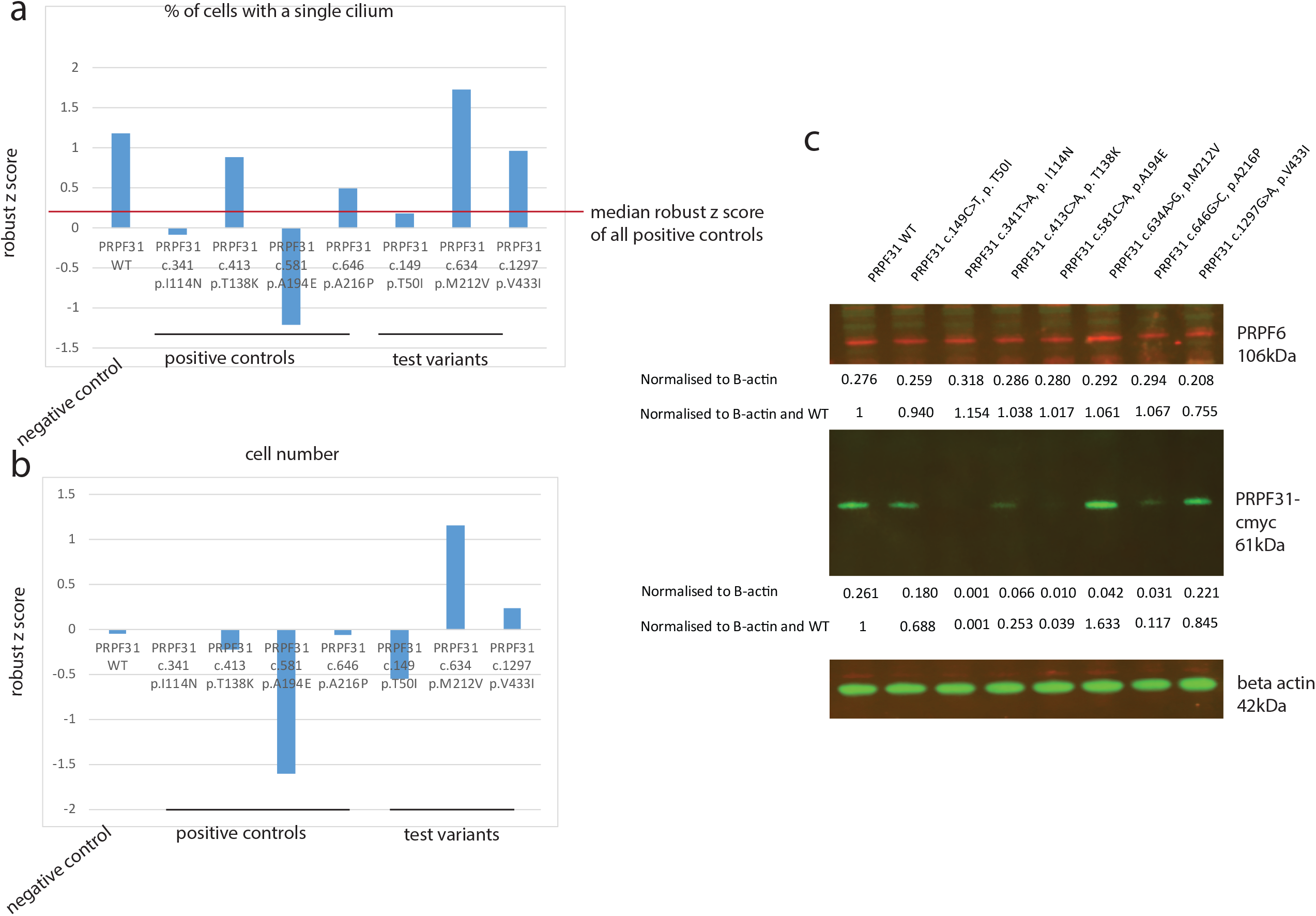
High throughput screening of the effect of expression of specific PRPF31 variants in *PRPF31*^*+/-*^ mutant clones. (a) Graph showing effect of expression of specific PRPF31 variants on the percentage of cells with a single cilium in *PRPF31*^*+/-*^ mutant clones. Effect is measured by robust z score, and median of all positive controls’ robust z scores is shown by a red line across the graph. (b) Graph showing effect of expression of specific PRPF31 variants on cell number in *PRPF31*^*+/-*^ mutant clones. (c) western blot showing level of expression of soluble PRPF6 (top), PRPF31 (middle) and beta-actin loading control (bottom). Intensity of bands are expressed normalized to beta-actin loading control and wild-type control.

Of the novel missenses being tested, PRPF31 c.149C>T p.Thr50Ile failed to fully rescue the defective cilia phenotype, suggesting that this variant is pathogenic (**Figure 4a**). PRPF31 c.634A>G p.Met212Val fully rescued the defective cilia phenotype, suggesting that this missense is benign (**Figure 4a**). PRPF31 c.1297G>A p.Val433Ile partially rescued the loss of cilia phenotype, suggesting that this may be a mildly pathogenic variant, similar to c.413C>A p.Thr138Lys, although we would more confidently ascribe this benign status (**Figure 4a**). A study of cell number showed that two of the missenses which showed the most severe impact on cilia (PRPF31 c.149C>T p.Thr50Ile and c.581C>A p.Ala194Glu) also caused a reduction in cell number, although c.341C>A p.Thr114Lys which is also a severe variant, does not (**Figure 4b**). However, overall there was no clear correlation between severity of effect on cilia phenotype and effect on cell number.

To confirm expression of each construct we extracted protein from transfected cells and analysed expression levels by western blotting. This showed that all constructs were expressed but, as previously reported (Wheway et al, 2019), some missense mutated forms of PRPF31 (c.341T>A p.Ile114Asn, c.581C>A p.Ala194Glu) were associated with reduced stability and solubility of the protein, appearing as lower levels in the soluble fraction of cell extracts (**Figure 4c**). The variants with the least soluble expression tended to be those with the most severe effect on cilia phenotype (**Figure 4c**). 3D structural analysis predicted that c.149C>T pT50I would interfere with binding to PRPF6. We did see a small decrease in total level of PRPF6 in cell transfected with this construct (**Figure 4c**) but did not investigate PRPF31/PRPF6 interactions.

Brnich et al 2019 suggest that with 10 or less control variants the maximum weight of evidence this assay can currently provide is BS3_supporting evidence (PRPF31 c.634A>G p.Met212Val, c.1297G>A p.Val433Ile) and PS3_supporting evidence (PRPF31 c.149C>T p.Thr50Ile). Using the OddsPath calculations of Brnich et al. 2019, with 5 classified control variants (4 pathogenic, 1 benign) with 1 intermediate result, the results of this assay provide BS3_supporting evidence that PRPF31 c.634A>G p.Met212Val and c.1297G>A p.Val433Ile are benign variants but indeterminate evidence that PRPF31 c.149C>T p.Thr50Ile is a pathogenic variant. To reach PS3_supporting evidence, we would need to include 2 additional confirmed benign variants, to reach PS3_moderate evidence we must include 4 additional benign variants.

## Discussion

Here we present a high-throughput high content imaging assay providing quantitative measure of effect of missense variants in the second most common cause of autosomal dominant RP, *PRPF31*. Our screening assay meets the criteria for a baseline standard *in vitro* test for clinical variant interpretation (Brnich et al. 2019) because the disease mechanism is understood (combined haploinsufficiency/dominant negative effects), the assay is applicable to this disease and this disease mechanism, normal/negative/wild-type (1) and abnormal/positive/null controls (4) are used on each assay plate, multiple replicates are used (each variant and control in 3 wells per plate, each plate repeated twice), variant controls are known benign and known pathogenic, and statistical analysis has been applied to calculate the level of evidence for each variant (robust z scores, OddsPath). This assay utilizes a new and well-characterized *PRPF31*^*+/-*^ human retinal cell line generated using CRISPR gene editing, which allows testing of *PRPF31* variants which may be causing disease through either haploinsufficiency or dominant negative effects, or a combination of both. The mutant cells have significantly fewer cilia than wild-type cells, allowing rescue of ciliogenesis with benign or mild variants, but do not totally lack cilia, so dominant negative effects can be observed.

The results of the assay provide BS3_supporting evidence to the benign classification of two novel uncharacterized *PRPF31* variants (c.634A>G pM212V and c.1297G>A p.V433I). The results of the assay also suggest that one novel uncharacterized *PRPF31* variant (c.149C>T pT50I) may be pathogenic, although 2 more controls would need to be included in the assay to allow this result to be used as PS3_supporting evidence. According to ACMG guidelines for clinical variant interpretation, PS3_supporting evidence in combination with other evidence of pathogenicity can allow a sequence variant to be classified as pathogenic or likely pathogenic (Richards et al. 2015). With the addition of 4 more control benign variants added to our assay, we can increase the weight of evidence provided by this assay to PS3_moderate, and we intend to develop our assay in this manner.

Providing *in vitro* evidence to aid classification of clinical variants is of significant importance to allow accurate genetic diagnoses to be made, to enable targeted testing of other family members, aid family planning, allow pre-implantation diagnosis and inform eligibility for gene therapy trials. With PRPF31 gene therapy in development, there is an urgent need for tools for accurate molecular diagnosis (Brydon et al. 2019).

The imaging-based screen uses a simple and robust image analysis algorithm to test a consistent cellular phenotype observed in *PRPF31* mutant cells; reduction in the number of cells with a single cilium. The assay provides a continuous data readout in the form of percentage of cells with a single cilium, which has the potential to provide more than a simple binary readout of pathogenic/benign but a measure of the extent of pathogenicity of each variant. The findings of this assay and other such assays can also provide novel insights into disease mechanism and prognosis. Whilst some variants simply fail to rescue loss of cilia in mutant cell lines and can be assumed to be causing disease by haploinsufficiency, others appear to have a dominant negative effect on cells. Those having dominant negative effects on cells appear to show the lowest levels of soluble protein expression. Although data relating to genotype-phenotype correlations in cases of patients with missense variants in *PRPF31* is sparse (Wheway et al. 2020), we hypothesise that the variants with the most significant effect on cilia will be associated with the earliest onset and worst prognosis.

Finally, our findings correlate to some extent with *in silico* predictions, although not perfectly. PolyPhen2 predicted c.149C>T pT50I to be benign, whereas our *in vitro* assay suggests this is a pathogenic variant. In this case, 3D structural analysis was additionally useful in predicting pathogenicity of this variant. However, neither CADD, SIFT, Polyphen nor 3D structural analyses predicted PRPF31 c.634A>G p.Met212Val to be benign, whereas our *in vitro* assay suggests this is benign. We suggest that these *in silico* tools are useful to identify most likely pathogenic variants, but that it is important to validate these predictions with *in vitro* assays.

## Data Availability

RNA sequence data is available via the Sequence Read Archive, accession PRJNA622794

https://www.ncbi.nlm.nih.gov/sra/PRJNA622794

## Funding

This work was supported by National Eye Research Centre Small Award SAC019, Wellcome Trust Seed Award in Science 204378/Z/16/Z, UWE Bristol Quality Research funds and University of Southampton Faculty of Medicine Research Management Committee funds.

## Acknowledgements

The authors would like to thank Dr Stephen Cross for assistance in high throughput imaging and analysis; Dr Carolann McGuire and Dr Richard Jewell for assistance in cell sorting. The authors acknowledge the use of the IRIDIS High Performance Computing Facility, and associated support services at the University of Southampton, in the completion of this work.

## Figure and Table Legends

**Table 1 - Summary of missense variants in PRPF31**

Published and ClinVar-deposited *PRPF31* variants predicted to cause missense changes in PRPF31 protein. The table summarises location, effect on splicing predicted by Human Splicing Finder, functional effect predicted by SIFT, PolyPhen2 and PyMol, and overall predicted pathogenicity. In green are highlighted positive control variants for our assay, in orange are highlighted variants to be tested.

**Supplementary Table 1**

Table showing sequence of sgRNA1 which was used to introduce CRISPR indel, genomic DNA sequence of potential off-target mapping sites, with 3 mismatches allowed (mismatches shown in lower case), chromosomal location of these potential off-target sites, whether they are on the + or – strand, number of mismatches, gene name and feature targeted.

**Supplementary Table 2**

Summary of analysis of potential off-target CRISPR cut sites, showing findings observed in IGV, through differential gene expression analysis by edgeR, and differential splicing analysis by rMATS, including alternative 3’ splice site usage (A3SS), alternative 5’ splice site usage (A5SS), mutually exclusive exons (MXE), retained introns (RI) and spliced exons (SE).

**Supplementary Figure 1 – Differential splicing of *PRPF31* intron 12-13 in wild-type and *PRPF31***^***+/-***^ **mutant clones**

(a) Sashimi plot showing statistically significantly lower levels of splicing of intron 12-13 in the nuclear RNA of PRPF31+/- clones compared to wild-type clones. (b) rMATS statistical analysis of this differential splicing in nucleus, showing intron inclusion level for wild-type and mutant clones, intron inclusion level difference and p values with a without correction for false discovery rate (FDR). (c) Sashimi plot showing no statistically significantly different level of inclusion of intron 12-13 in the cytoplasmic RNA of PRPF31^+/-^ clones compared to wild-type clones. (d) rMATS statistical analysis of this differential splicing in cytoplasm, showing intron inclusion level for wild-type and mutant clones, intron inclusion level difference and p values with a without correction for false discovery rate (FDR). SJ = only reads mapping to splice junctions considered SJ + I = reads mapping to splice junctions and to intron considered.

## Notes

### Competing Interest Statement

The authors have declared no competing interest.

## References

Adzhubei I, Jordan DM, Sunyaev SR. 2013. Predicting functional effect of human missense mutations using PolyPhen-2. Curr Protoc Hum Genet Chapter 7: Unit7.20.

Bachmann-Gagescu R, Dempsey JC, Phelps IG, O’Roak BJ, Knutzen DM, Rue TC, Ishak GE, Isabella CR, Gorden N, Adkins J et al. 2015. Joubert syndrome: a model for untangling recessive disorders with extreme genetic heterogeneity. Journal of medical genetics 52: 514–522.

Bae S, Park J, Kim JS. 2014. Cas-OFFinder: a fast and versatile algorithm that searches for potential off-target sites of Cas9 RNA-guided endonucleases. Bioinformatics 30: 1473–1475.

Bertram K, Agafonov DE, Dybkov O, Haselbach D, Leelaram MN, Will CL, Urlaub H, Kastner B, Luhrmann R, Stark H. 2017. Cryo-EM Structure of a Pre-catalytic Human Spliceosome Primed for Activation. Cell 170: 701-713.e711.

Birmingham A, Selfors LM, Forster T, Wrobel D, Kennedy CJ, Shanks E, Santoyo-Lopez J, Dunican DJ, Long A, Kelleher D et al. 2009. Statistical methods for analysis of high-throughput RNA interference screens. Nat Meth 6: 569–575.

Brnich SE, Abou Tayoun AN, Couch FJ, Cutting GR, Greenblatt MS, Heinen CD, Kanavy DM, Luo X, McNulty SM, Starita LM et al. 2019. Recommendations for application of the functional evidence PS3/BS3 criterion using the ACMG/AMP sequence variant interpretation framework. Genome Med 12: 3.

Brydon EM, Bronstein R, Buskin A, Lako M, Pierce EA, Fernandez-Godino R. 2019. AAV-Mediated Gene Augmentation Therapy Restores Critical Functions in Mutant PRPF31. Mol Ther Methods Clin Dev 15: 392–402.

Buskin A, Zhu L, Chichagova V, Basu B, Mozaffari-Jovin S, Dolan D, Droop A, Collin J, Bronstein R, Mehrotra S et al. 2018. Disrupted alternative splicing for genes implicated in splicing and ciliogenesis causes PRPF31 retinitis pigmentosa. Nat Commun 9: 4234.

Carpenter AE, Jones TR, Lamprecht MR, Clarke C, Kang IH, Friman O, Guertin DA, Chang JH, Lindquist RA, Moffat J et al. 2006. CellProfiler: image analysis software for identifying and quantifying cell phenotypes. Genome biology 7: R100-2006-2007-2010-r2100. Epub 2006 Oct 2031.

Chung N, Zhang XD, Kreamer A, Locco L, Kuan P-F, Bartz S, Linsley PS, Ferrer M, Strulovici B. 2008. Median Absolute Deviation to Improve Hit Selection for Genome-Scale RNAi Screens. Journal of Biomolecular Screening 13: 149–158.

Desmet FO, Hamroun D, Lalande M, Collod-Beroud G, Claustres M, Beroud C. 2009. Human Splicing Finder: an online bioinformatics tool to predict splicing signals. Nucleic acids research 37: e67.

Ferrante MI, Giorgio G, Feather SA, Bulfone A, Wright V, Ghiani M, Selicorni A, Gammaro L, Scolari F, Woolf AS et al. 2001. Identification of the gene for oral-facial-digital type I syndrome. Am J Hum Genet 68: 569–576.

Frankish A, Diekhans M, Ferreira AM, Johnson R, Jungreis I, Loveland J, Mudge JM, Sisu C, Wright J, Armstrong J et al. 2019. GENCODE reference annotation for the human and mouse genomes. Nucleic Acids Res 47: D766–D773.

Huang da W, Sherman BT, Lempicki RA. 2009. Bioinformatics enrichment tools: paths toward the comprehensive functional analysis of large gene lists. Nucleic acids research 37: 1–13.

Kanavy DM, McNulty SM, Jairath MK, Brnich SE, Bizon C, Powell BC, Berg JS. 2019. Comparative analysis of functional assay evidence use by ClinGen Variant Curation Expert Panels. Genome Med 11: 77.

Kim J, Lee JE, Heynen-Genel S, Suyama E, Ono K, Lee K, Ideker T, Aza-Blanc P, Gleeson JG. 2010. Functional genomic screen for modulators of ciliogenesis and cilium length. Nature 464: 1048–U1114.

Kim JH, Ki SM, Joung JG, Scott E, Heynen-Genel S, Aza-Blanc P, Kwon CH, Kim J, Gleeson JG, Lee JE. 2016. Genome-wide screen identifies novel machineries required for both ciliogenesis and cell cycle arrest upon serum starvation. Biochimica et biophysica acta 1863: 1307–1318.

Knopp C, Rudnik-Schoneborn S, Eggermann T, Bergmann C, Begemann M, Schoner K, Zerres K, Ortiz Bruchle N. 2015. Syndromic ciliopathies: From single gene to multi gene analysis by SNP arrays and next generation sequencing. Molecular and cellular probes 29: 299–307.

Landrum MJ, Lee JM, Benson M, Brown G, Chao C, Chitipiralla S, Gu B, Hart J, Hoffman D, Hoover J et al. 2016. ClinVar: public archive of interpretations of clinically relevant variants. Nucleic acids research 44: D862–868.

Landrum MJ, Lee JM, Riley GR, Jang W, Rubinstein WS, Church DM, Maglott DR. 2014. ClinVar: public archive of relationships among sequence variation and human phenotype. Nucleic acids research 42: D980–985.

Liu S, Li P, Dybkov O, Nottrott S, Hartmuth K, Luhrmann R, Carlomagno T, Wahl MC. 2007. Binding of the human Prp31 Nop domain to a composite RNA-protein platform in U4 snRNP. Science (New York, NY) 316: 115–120.

Martin-Merida I, Aguilera-Garcia D, Fernandez-San Jose P, Blanco-Kelly F, Zurita O, Almoguera B, Garcia-Sandoval B, Avila-Fernandez A, Arteche A, Minguez P et al. 2018. Toward the Mutational Landscape of Autosomal Dominant Retinitis Pigmentosa: A Comprehensive Analysis of 258 Spanish Families. Investigative ophthalmology & visual science 59: 2345–2354.

McCarthy DJ, Chen Y, Smyth GK. 2012. Differential expression analysis of multifactor RNA-Seq experiments with respect to biological variation. Nucleic Acids Res 40: 4288–4297.

McLaren W, Gil L, Hunt SE, Riat HS, Ritchie GR, Thormann A, Flicek P, Cunningham F. 2016. The Ensembl Variant Effect Predictor. Genome Biol 17: 122.

Ng PC, Henikoff S. 2003. SIFT: Predicting amino acid changes that affect protein function. Nucleic Acids Res 31: 3812–3814.

Oud MM, Lamers IJ, Arts HH. 2017. Ciliopathies: Genetics in Pediatric Medicine. Journal of pediatric genetics 6: 18–29.

Rentzsch P, Witten D, Cooper GM, Shendure J, Kircher M. 2019. CADD: predicting the deleteriousness of variants throughout the human genome. Nucleic Acids Res 47: D886–D894.

Richards S, Aziz N, Bale S, Bick D, Das S, Gastier-Foster J, Grody WW, Hegde M, Lyon E, Spector E et al. 2015. Standards and guidelines for the interpretation of sequence variants: a joint consensus recommendation of the American College of Medical Genetics and Genomics and the Association for Molecular Pathology. Genet Med 17: 405–424.

Robinson JT, Thorvaldsdóttir H, Winckler W, Guttman M, Lander ES, Getz G, Mesirov JP. 2011. Integrative genomics viewer. Nat Biotechnol 29: 24–26.

Robinson MD, McCarthy DJ, Smyth GK. 2010. edgeR: a Bioconductor package for differential expression analysis of digital gene expression data. Bioinformatics 26: 139–140.

Roosing S, Hofree M, Kim S, Scott E, Copeland B, Romani M, Silhavy JL, Rosti RO, Schroth J, Mazza T et al. 2015. Functional genome-wide siRNA screen identifies KIAA0586 as mutated in Joubert syndrome. Elife 4: e06602.

Rose AM, Bhattacharya SS. 2016. Variant haploinsufficiency and phenotypic non-penetrance in PRPF31-associated retinitis pigmentosa. Clin Genet 90: 118–126.

Sawyer SL, Hartley T, Dyment DA, Beaulieu CL, Schwartzentruber J, Smith A, Bedford HM, Bernard G, Bernier FP, Brais B et al. 2016. Utility of whole-exome sequencing for those near the end of the diagnostic odyssey: time to address gaps in care. Clinical genetics 89: 275–284.

Shen S, Park JW, Lu ZX, Lin L, Henry MD, Wu YN, Zhou Q, Xing Y. 2014. rMATS: robust and flexible detection of differential alternative splicing from replicate RNA-Seq data. Proc Natl Acad Sci U S A 111: E5593–5601.

Watson CM, Crinnion LA, Berry IR, Harrison SM, Lascelles C, Antanaviciute A, Charlton RS, Dobbie A, Carr IM, Bonthron DT. 2016. Enhanced diagnostic yield in Meckel-Gruber and Joubert syndrome through exome sequencing supplemented with split-read mapping. BMC Medical Genetics 17: 1.

Wheway G, Douglas A, Baralle D, Guillot E. 2020. Mutation spectrum of PRPF31, genotype-phenotype correlation in retinitis pigmentosa, and opportunities for therapy. Exp Eye Res 192: 107950.

Wheway G, Lord J, Baralle D. 2019a. Splicing in the pathogenesis, diagnosis and treatment of ciliopathies. Biochim Biophys Acta Gene Regul Mech 1862: 194433.

Wheway G, Mitchison HM. 2019. Opportunities and Challenges for Molecular Understanding of Ciliopathies-The 100,000 Genomes Project. Frontiers in genetics 10: 127.

Wheway G, Nazlamova L, Meshad N, Hunt S, Jackson N, Churchill A. 2019b. A Combined in silico, in vitro and Clinical Approach to Characterize Novel Pathogenic Missense Variants in PRPF31 in Retinitis Pigmentosa. Frontiers in genetics 10: 248.

Wheway G, Schmidts M, Mans DA, Szymanska K, Nguyen TM, Racher H, Phelps IG, Toedt G, Kennedy J, Wunderlich KA et al. 2015. An siRNA-based functional genomics screen for the identification of regulators of ciliogenesis and ciliopathy genes. Nature cell biology 17: 1074–1087.

Xiao X, Cao Y, Zhang Z, Xu Y, Zheng Y, Chen LJ, Pang CP, Chen H. 2017. Novel Mutations in PRPF31 Causing Retinitis Pigmentosa Identified Using Whole-Exome Sequencing. Investigative ophthalmology & visual science 58: 6342–6350.

Zhang XD. 2007. A pair of new statistical parameters for quality control in RNA interference high-throughput screening assays. Genomics 89: 552–561.

